# Targeted Gene Sequencing for Head and Neck Cancer Candidate Genes in the Nepal case-control study

**DOI:** 10.64898/2026.10.27.26366653

**Authors:** Ishor Thapa, Chun-Pin Esther Chang, Angela K Snow, Judith Rosenthal, Russell Bell, Austin Wood, Bhola Siwakoti, Dej Kumar Gautam, Amir Sapkota, Sean V Tavtigian, Bing-Jian Feng, Mia Hashibe

**Affiliations:** Department of Oncological Sciences, University of Utah School of Medicine, Salt Lake City, UT 84112, USA; Huntsman Cancer Institute at the University of Utah, Salt Lake City, UT 84112, USA; Department of Family Medicine & Public Health, University of Utah School of Medicine, Salt Lake City, UT 84108, USA; Department of Cancer Prevention, Control and Research, B.P. Koirala Memorial Cancer Hospital, Chitwan, Bharatpur 44200, Nepal; Department of Surgical Oncology, B.P. Koirala Memorial Cancer Hospital, Chitwan, Bharatpur 44200, Nepal; Department of Epidemiology and Biostatistics, University of Maryland School of Public Health, College Park, MD 20740, USA; Department of Dermatology, University of Utah School of Medicine, Salt Lake City, UT 84103, USA; Department of Epidemiology, UCLA Fielding School of Public Health, Los Angeles, CA 90095, USA

**Keywords:** head and neck cancer, targeted gene sequencing, cancer predisposition genes, genetic epidemiology

## Abstract

Head and Neck cancer (HNC) remains a major global cancer burden, with high incidence and mortality in low- and middle-income countries. Although several genetic variants associated with HNC risk have been identified, cancer predisposition genes in underrepresented populations remain poorly understood. We conducted a hospital-based case-control study at B.P. Koirala Memorial Cancer Hospital in Nepal with 315 HNC cases and 323 controls. Demographic, lifestyle, and clinical data were collected through in-person interviews. Targeted gene sequencing of 493 candidate HNC risk genes were performed, followed by gene- and pathway-based association analyses, and clinical annotation of the identified variants. Consistent with prior findings, HNC incidence increased significantly with alcohol and cigarette use, lower education, and lower socio-economic status. Gene-based analyses identified several genes with suggestive associations prior to multiple-testing adjustment, including *HSPG2*, *KRT6B*, *SLC4A2*, and *FREM1*. Similarly, no pathways remained statistically significant after adjustment. However, it identified potential biological processes, including extracellular matrix organization, epithelial differentiation, and transcription regulation. Analysis of six cancer predisposition genes (*APC*, *BAP1*, *BRCA1*, *BRCA2*, *RAD51B*, and *MUTYH*) identified pathogenic and likely pathogenic variants with clinical relevance, although no significant enrichment was observed between cases and controls. This study provides the first targeted sequencing analysis combined with epidemiological data for HNC in the Nepalese population, an underrepresented high-risk group. While no significant genetic associations were identified, clinically relevant rare variants were detected. Larger studies in South Asian populations are needed to improve the detection of rare variants and to inform risk assessment.

**Novelty and Impact:** This study presents the first integrated targeted sequencing and epidemiological analysis of head and neck cancer in the Nepalese population, an underrepresented and understudied high-risk group. Using a validated 493-gene panel, we sequenced 315 HNC cases and 323 controls. We identified clinically relevant pathogenic and likely pathogenic variants in known susceptibility genes, including *BRCA1*, *BRCA2*, *RAD51B*, and *MUTYH*. This study also confirmed the strong associations of tobacco, alcohol, and chewing habits with HNC risk, emphasizing the need to study cancer risk in different populations and cultural settings.

## Introduction

Head and Neck Cancers (HNC) collectively include cancers of the oral cavity, oropharynx, hypopharynx, and larynx.^1^ Globally, HNC ranked as the sixth most prevalent cancer in 2024 as per Global Cancer Statistics, with over 845,000 new diagnoses and more than 454,000 deaths annually.^2^ This represents 4.6% of all cancer diagnoses and 4.5% of cancer deaths.

Previous studies have identified several important environmental risk factors for HNC. Alcohol and tobacco use are major risk factors. There is a synergistic effect whereas their combined use significantly increases the HNC risk over that of alcohol or tobacco use alone, the magnitude of which depends on the duration and frequency of exposure.^3^ Human Papillomavirus (HPV) infection is also a strong risk factor, especially for oropharyngeal cancers.^4^ Other less-reported risk factors include household air pollution, occupational and radiation exposure.^5,6^

Studies on genetic risk factors for HNC provide evidence that pathways involved in DNA damage repair, cell-cycle regulation, apoptosis, and immune modulation contribute to HNC susceptibility.^7^ Genome-wide association studies (GWAS) across diverse populations have identified several HNC susceptibility loci. The Million Veteran Program reported associations of the *DMBT1* and *TPM1* loci with oral cavity, hypopharyngeal, and laryngeal cancers, and of the *CRTAM*, *CDH5*, and *CDH2* loci with oropharyngeal cancer.^8^ In the same study, variants associated with HNC risk in African-American patients included loci in immune-related genes such as *HLA-G*, *GAS1*, *NCAM1*, *CD6*, and *CD68*. A study of European populations found associations near alcohol-metabolism genes (*ADH* clusters, *ALDH2*) and the DNA repair gene *HEL308*.^9^ Further, studies in East Asian populations identified *FADS1*, *AIF1*, *TBX5*, *HLA-B/DQ*, *TERT-CLMPT1L*, *ADH1B*, *LAMC3*, *SKIV2L*, and *TNXB* as risk loci specifically in laryngeal squamous cell carcinoma.^10,11^ However, no high-penetrance predisposition gene specific to HNC has been fully established.

A recent global trend shows a concerning increase in HNC cases in low- and middle-income countries (LMIC), which now accounts for two-thirds of global HNC cases.^2^ Notably, Asia, particularly South Asia, reports a disproportionately high burden of Head and Neck Squamous Cell carcinoma (HNSCC) compared to its general population.^12^ Interestingly, studies in the United States and Canada have demonstrated a higher rate of HNSCC among South Asian diaspora populations than in the general populations of those countries.^13,14^ Higher incidence in developing countries is largely attributed to the widespread use of alcohol, tobacco, and smokeless tobacco products like betel quid, which is compounded by poor oral hygiene and limited health resources for prevention and treatment.^4,15–18^ Given the significant variation in HNC etiology across genetic backgrounds and geographical regions, gaining a comprehensive understanding of its global impact remains challenging.^19,20^

The present case-control study focuses on Nepal, a LMIC where HNC is becoming a major public health issue.^21^ A previous case-control study in Nepal, involving 549 HNC cases and 601 controls, confirmed a significant increase in HNC risk associated with tobacco smoking, chewing (Pan, Khaini, Surti, Zarda, Pan masala, and Supari), and/or alcohol consumption.^17,22^ Exposure to all three risk factors led to a 13-fold increase in HNC risk.^17^ The present study, the first of its kind in Nepal, builds upon these findings by integrating demographic and lifestyle information with phenotypic data and genotypic data from targeted exome sequencing.

## Materials and Methods

### Study population

A hospital-based case-control study was conducted at B.P. Koirala Memorial Cancer Hospital (BPKMCH) in Chitwan district, Nepal (Figure 1). BPKMCH is the National Cancer Center and the main cancer referral center in Nepal. From 2016 to 2018, we recruited HNC cases and frequency-matched controls based on age (±5 years), sex, race/ethnicity, and residence. The inclusion criteria for HNC cases were: (1) being a patient at BPKMCH; (2) having resided in Nepal for a minimum of five years; and (3) being at least 18 years old. The sub-sites of HNC included the oral cavity, oropharynx, hypopharynx, and larynx. Cases involving the salivary gland (including parotid glands), nasopharynx, and thyroid were excluded. Incident cancer cases were recruited within three months of the initial diagnosis. False-positive cases were excluded following the final pathological diagnosis. The inclusion criteria for hospital-based controls were being cancer-free, aged 18 or older, and visiting BPKMCH. The reasons for visiting the hospital included cancer screening (22.0%), being a hospital visitor (63.6%), or both (14.5%). Individuals for cases and controls were recruited from April 2016 to July 2018. Blood sample collections and in-person interviews were conducted by trained staff members. Information on demographic factors, occupational history, dietary habits, medical history, residential history, reproductive history (for women only), quality of life (for cases only), and detailed histories of tobacco smoking, chewing habits, and alcohol consumption were collected using a questionnaire.

**Figure 1:**
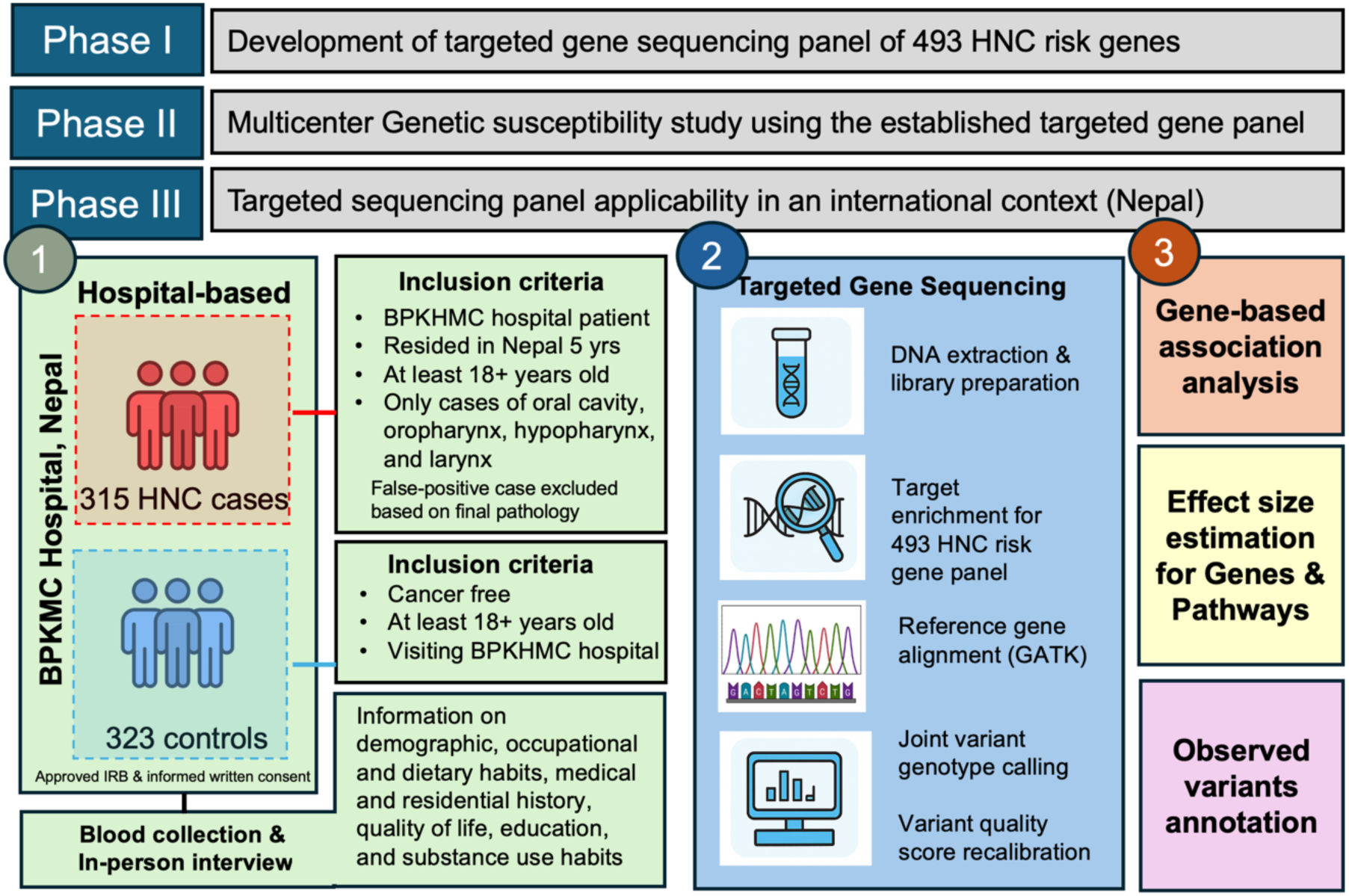
Schematics of overall study design and workflow. Phase I focused on developing and validating a targeted sequencing panel of 493 HNC risk genes. Phase II and Phase III evaluates the utility of the gene sequencing panel in an international context, with Phase III focused on Nepal. (1) At BPKHM Hospital in Nepal, a case-control study was conducted enrolling 315 HNC cases and 323 controls with defined inclusion criteria, blood collection, and in-person interviews. (2) Targeted gene sequencing included DNA extraction, library preparation & target enrichment, sequencing, reference-based alignment, joint variant genotype calling, and variant quality score recalibration. (3) Gene- and pathway-based association analyses were performed, followed by clinical and functional annotation of the observed variants.

### Targeted gene sequencing of Nepal samples

Briefly, genomic DNA was extracted from peripheral blood samples using the Gentra Puregene Blood Kit (Qiagen #158467). DNA libraries were prepared from samples with a 260/280 ≥ 1.4 and concentration >20 ng/µL using the Twist Library Preparation EF Kit (Twist Bioscience #104119) as per manufacture protocol. Target enrichment was performed prior to sequencing using the Twist Standard Hybridization Reagent Kit v2 (Twist Bioscience #101026). A custom targeted sequencing panel (Twist Bioscience #101004) of 493 genes comprising known cancer susceptibility genes and prospective HNC risk genes from analysis of a prior study was used in the target enrichment step.^23^ Sequencing was performed at the HCI High-Throughput Genomic Core Facility. The GATK best practices workflow was applied for reference genome alignment, joint variant genotype calling, and variant quality score recalibration.^24^ The sequencing reads were mapped to the human reference genome (HG38) using Burrows-Wheeler Aligner BWA v0.7.9a.^25,26^ Sequence runs with 95% of target bases with ≥ 20X and 90% ≥ 30X coverage, as determined by GATK’s CollectHsMetrics, were advanced to the next step. Duplicate reads were marked with Biobambam2 sortmadup.^27^ Local realignment was achieved using GATK’s IndelRealigner, and base quality score was adjusted with GATK’s BaseRecalibrator. Only uniquely mapped reads with mapping quality ≥ 30 were retained. Per-sample gVCF files from GATK’s HaplotypeCaller were jointly genotyped using GATK’s GenotypeGVCFs.

### Gene-based association analysis

We performed gene-based association tests using the Variant Interpretation for Clinical Testing Or Research (VICTOR) package on variants with a minor allele frequency (MAF) < 0.01.^28^ We conducted Firth’s logistic regression on two sets of variants: loss-of-function (LoF) variants alone and a combination of LoF and predicted damaging variants. The BayesDel score was used to assess the damaging effect of each variant.^29^ Functional annotations were based on the RefSeq database, restricted to principal transcripts defined by the APPRIS database.^30^ LoF variants were defined as frameshift, stop-gain, and splice-site variants, excluding those located in the last 5% of the coding region. Splice-site variants were defined as those within two base pairs of an intron. For other variants near splice donor (−3 to +8 bp) or acceptor (−12 to +2 bp) sites, splicing impact was assessed using MaxEntScan.^28^ Variants classified as benign in the ClinVar database were excluded from the analysis. All gene-based analyses were adjusted for age, sex, and the first three principal components to account for population structure and technical variation.

### Effect size estimation for genes and pathways

We evaluated variant effect sizes of the top genes in association tests for head and neck cancer risk across a variety of functional classifications, including amino acid changes, in silico functional predictions, and ClinVar annotation status, using ANNOVAR (Version 20221231).^29^ For ClinVar annotations, we included ‘pathogenic’ and ‘likely pathogenic’ variants in the known pathogenic category, and uncertain or unknown significance as VUS. The effect size was estimated using the Firth regression method for each population group. We used the first ten PCs, age, and sex as covariates. To avoid bias due to the small sample size, we required at least 5 carriers of variants in cases and controls to estimate the odds ratio. Pathway-based analysis was conducted using Gene Set Enrichment Analysis to search MSigDB following guidelines outlined in the VICTOR documentation.^28,30,31^

### Computation tool for variant effect prediction and visualization

To predict the effect of missense substitution on protein function, we used AlphaMissense^32^ and BayesDel.^29^ To visualize the location of various missense substitutions across the protein domains as a lollipop diagram, the web version of SRplot was used.^33^

## Results

The analysis included 315 HNC cases and 323 controls (Figure 1). Among HNC cases, more than 60% of the cases had oral cancer, followed by laryngeal (15.9%), oropharyngeal (10.5%), and hypopharyngeal (9.2%) cancers. Controls were matched to HNC cases on age, sex, ethnicity, and residence; accordingly, no significant differences were observed in these variables between groups (Table 1). However, there were significant differences in lifestyle and socioeconomic status. Consistent with previous studies, cigarette smoking was significantly more common in cases (63.5%) than controls (45.2%) (p < 0.001). The prevalence of chewing habits was significantly higher among cases (66.7%) than controls (51.7%) (p < 0.001). Among cases (67%), alcohol consumption was more frequent than in controls (52.9%) (p < 0.001). Similarly, educational status was significantly lower with 51% of cases reporting no formal education compared to 33.4% of controls (p < 0.001). Additionally, family income was significantly lower in cases, with only 42.2% reporting a monthly income of ≥ 5000 NPR (Nepalese Rupee), versus 58.5% among controls (p < 0.001).

**Table 1.**
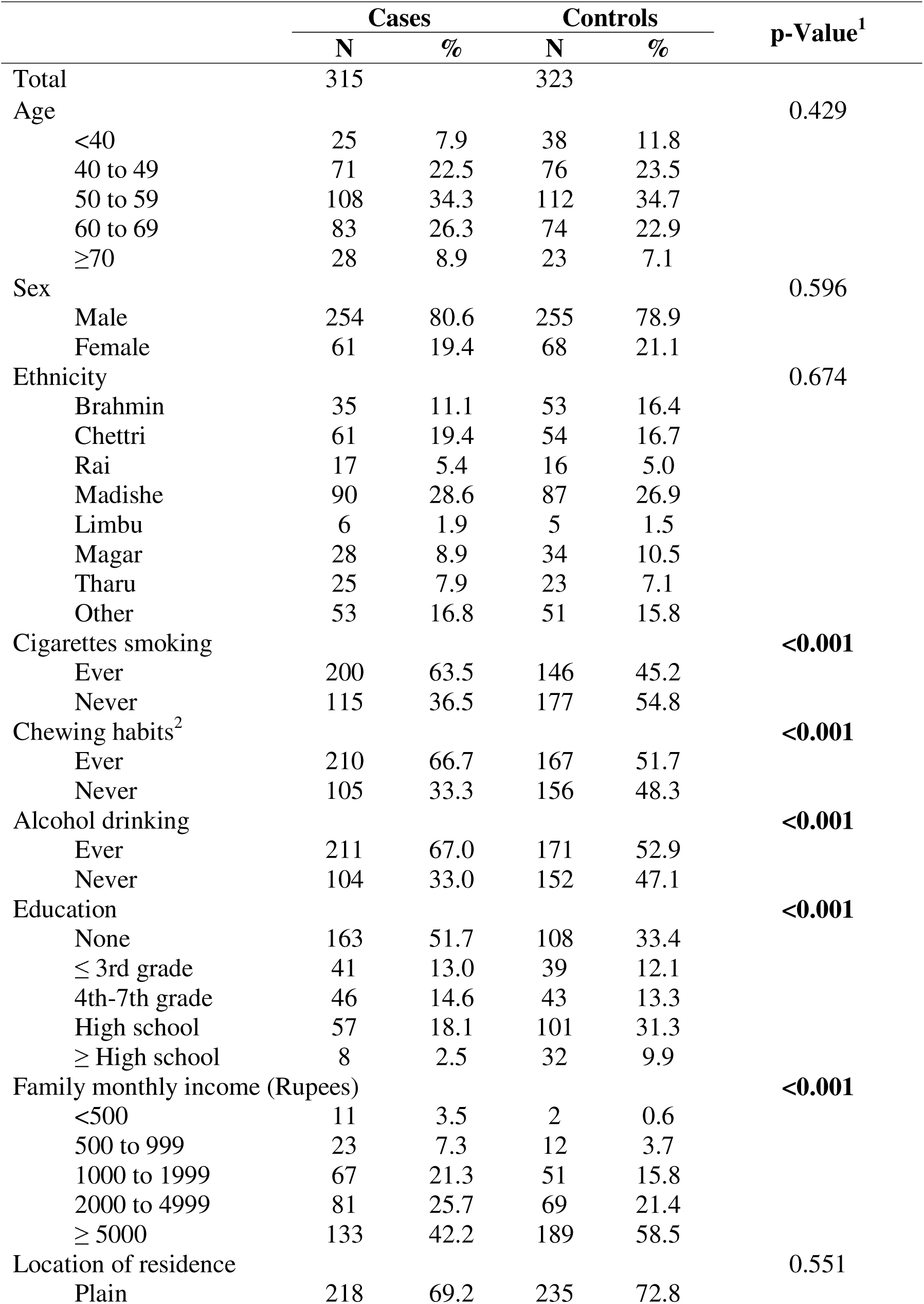
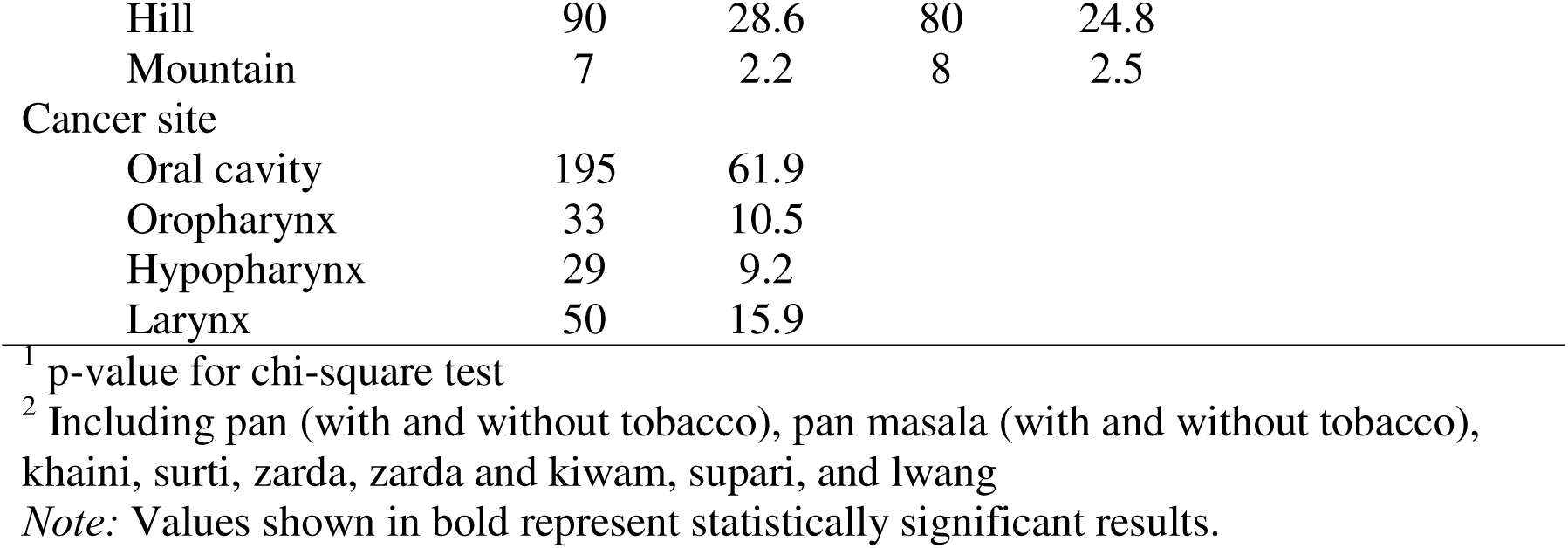
The distribution of selected characteristics of head and neck cancer cases and controls in the Nepal case-control study.

|  | Cases |  | Controls |  | p-Value<sup>1</sup> |
| --- | --- | --- | --- | --- | --- |
|  | N | % | N | % |  |
| Total | 315 |  | 323 |  |  |
| Age |  |  |  |  | 0.429 |
| <40 | 25 | 7.9 | 38 | 11.8 |  |
| 40 to 49 | 71 | 22.5 | 76 | 23.5 |  |
| 50 to 59 | 108 | 34.3 | 112 | 34.7 |  |
| 60 to 69 | 83 | 26.3 | 74 | 22.9 |  |
| ≥70 | 28 | 8.9 | 23 | 7.1 |  |
| Sex |  |  |  |  | 0.596 |
| Male | 254 | 80.6 | 255 | 78.9 |  |
| Female | 61 | 19.4 | 68 | 21.1 |  |
| Ethnicity |  |  |  |  | 0.674 |
| Brahmin | 35 | 11.1 | 53 | 16.4 |  |
| Chettri | 61 | 19.4 | 54 | 16.7 |  |
| Rai | 17 | 5.4 | 16 | 5.0 |  |
| Madishe | 90 | 28.6 | 87 | 26.9 |  |
| Limbu | 6 | 1.9 | 5 | 1.5 |  |
| Magar | 28 | 8.9 | 34 | 10.5 |  |
| Tharu | 25 | 7.9 | 23 | 7.1 |  |
| Other | 53 | 16.8 | 51 | 15.8 |  |
| Cigarettes smoking |  |  |  |  | <0.001 |
| Ever | 200 | 63.5 | 146 | 45.2 |  |
| Never | 115 | 36.5 | 177 | 54.8 |  |
| Chewing habits<sup>2</sup> |  |  |  |  | <0.001 |
| Ever | 210 | 66.7 | 167 | 51.7 |  |
| Never | 105 | 33.3 | 156 | 48.3 |  |
| Alcohol drinking |  |  |  |  | <0.001 |
| Ever | 211 | 67.0 | 171 | 52.9 |  |
| Never | 104 | 33.0 | 152 | 47.1 |  |
| Education |  |  |  |  | <0.001 |
| None | 163 | 51.7 | 108 | 33.4 |  |
| ≤ 3rd grade | 41 | 13.0 | 39 | 12.1 |  |
| 4th-7th grade | 46 | 14.6 | 43 | 13.3 |  |
| High school | 57 | 18.1 | 101 | 31.3 |  |
| ≥ High school | 8 | 2.5 | 32 | 9.9 |  |
| Family monthly income (Rupees) |  |  |  |  | <0.001 |
| <500 | 11 | 3.5 | 2 | 0.6 |  |
| 500 to 999 | 23 | 7.3 | 12 | 3.7 |  |
| 1000 to 1999 | 67 | 21.3 | 51 | 15.8 |  |
| 2000 to 4999 | 81 | 25.7 | 69 | 21.4 |  |
| ≥ 5000 | 133 | 42.2 | 189 | 58.5 |  |
| Location of residence |  |  |  |  | 0.551 |
| Plain | 218 | 69.2 | 235 | 72.8 |  |

|  |  |  |  |  |
| --- | --- | --- | --- | --- |
| Hill | 90 | 28.6 | 80 | 24.8 |
| Mountain | 7 | 2.2 | 8 | 2.5 |
| Cancer site |  |  |  |  |
| Oral cavity | 195 | 61.9 |  |  |
| Oropharynx | 33 | 10.5 |  |  |
| Hypopharynx | 29 | 9.2 |  |  |
| Larynx | 50 | 15.9 |  |  |

<sup>1</sup> p-value for chi-square test
<sup>2</sup> Including pan (with and without tobacco), pan masala (with and without tobacco), khaini, surti, zarda, zarda and kiwam, supari, and Iwang
*Note:* Values shown in bold represent statistically significant results.

Gene-based analysis identified associations between HNC risk and *HSPG2*, *KRT6B*, *SLC4A2*, and *FREM* (Table 2); however, after adjusting for multiple comparisons, these associations were not statistically significant. Similarly, in the pathway analysis, none of the associations were statistically significant after multiple-comparison adjustment. Nonetheless, ten pathways had a smaller nominal p-value than any individual gene. Theses ten pathways, ranked by p-value, are listed in Table 3. Among these, *HSPG2* is involved in nine pathways, including mammary gland development, extracellular matrix disassembly, stromal cell signaling, histone acetylation regulation, and Cyclin D1-associated genes.

**Table 2.** Gene-based analysis in the Nepal case-control study, top 20 genes.

| **Gene** | **Chromosome** | **OR [95% CI]** | **Nominal p-value\*** |
| --- | --- | --- | --- |
| *HSPG2* | 1p36.12 | 9.48 [1.94, 46.3] | 0.0027 |
| *KRT6B* | 12q13.13 | 11.5 [1.35, 97.4] | 0.013 |
| *SLC4A2* | 7q36.1 | 9.05 [1.24, 65.8] | 0.015 |
| *FREM1* | 9p22.3 | 2.15 [1.00, 4.62] | 0.025 |
| *ASPM* | 1q31.3 | 3.67 [0.75, 18.1] | 0.055 |
| *UBR5* | 8q22.3 | 3.33 [0.75, 14.8] | 0.057 |
| *AACS* | 12q24.31 | 1.60 [0.81, 3.14] | 0.087 |
| *SCARA3* | 8p21.1 | 3.51 [0.57, 21.7] | 0.088 |
| *POLL* | 10q24.32 | 1.90 [0.74, 4.85] | 0.09 |
| *RYR3* | 15q14 | 1.29 [0.89, 1.88] | 0.091 |
| *TSPAN15* | 10q22.1 | 1.72 [0.77, 3.88] | 0.094 |
| *KCNJ1* | 11q24.3 | 3.21 [0.51, 20.3] | 0.11 |
| *RAF1* | 3p25.2 | 2.49 [0.56, 11.0] | 0.11 |
| *ASNS* | 7q21.3 | 2.49 [0.42, 14.6] | 0.16 |
| *FANCM* | 14q21.2 | 1.68 [0.59, 4.83] | 0.17 |
| *PDHX* | 11p13 | 2.21 [0.44, 11.2] | 0.17 |
| *CHD1L* | 1q21.1 | 1.81 [0.47, 6.92] | 0.19 |
| *EDEM1* | 3p26.1 | 1.68 [0.51, 5.60] | 0.2 |
| *ALK* | 2p23.2-2p23.1 | 1.79 [0.46, 6.98] | 0.2 |
| *ACADL* | 2q34 | 1.45 [0.59, 3.56] | 0.21 |

\*No statistical significance after adjusting for multiple comparisons
OR: Odds ratio; CI: Confidence Interval

**Table 3.**
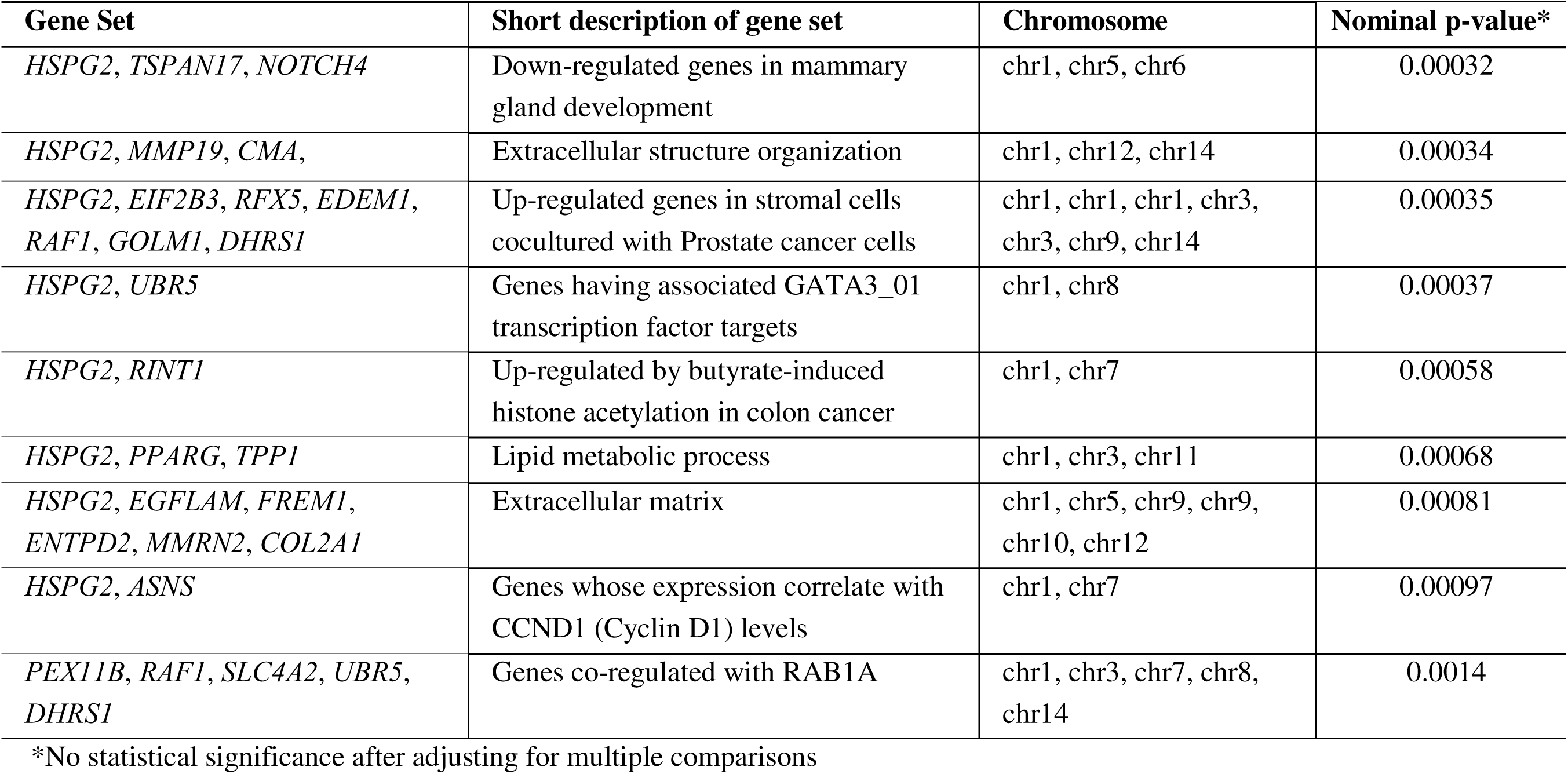
Pathway analysis in the Nepal case-control study.

| **Gene Set** | **Short description of gene set** | **Chromosome** | **Nominal p-value\*** |
| --- | --- | --- | --- |
| *HSPG2, TSPAN17, NOTCH4* | Down-regulated genes in mammary gland development | chr1, chr5, chr6 | 0.00032 |
| *HSPG2, MMP19, CMA,* | Extracellular structure organization | chr1, chr12, chr14 | 0.00034 |
| *HSPG2, EIF2B3, RFX5, EDEM1, RAF1, GOLM1, DHRS1* | Up-regulated genes in stromal cells cocultured with Prostate cancer cells | chr1, chr1, chr1, chr3, chr3, chr9, chr14 | 0.00035 |
| *HSPG2, UBR5* | Genes having associated GATA3_01 transcription factor targets | chr1, chr8 | 0.00037 |
| *HSPG2, RINT1* | Up-regulated by butyrate-induced histone acetylation in colon cancer | chr1, chr7 | 0.00058 |
| *HSPG2, PPARG, TPP1* | Lipid metabolic process | chr1, chr3, chr11 | 0.00068 |
| *HSPG2, EGFLAM, FREM1, ENTPD2, MMRN2, COL2A1* | Extracellular matrix | chr1, chr5, chr9, chr9, chr10, chr12 | 0.00081 |
| *HSPG2, ASNS* | Genes whose expression correlate with CCND1 (Cyclin D1) levels | chr1, chr7 | 0.00097 |
| *PEX11B, RAF1, SLC4A2, UBR5, DHRS1* | Genes co-regulated with RAB1A | chr1, chr3, chr7, chr8, chr14 | 0.0014 |

\*No statistical significance after adjusting for multiple comparisons

We further examined six genes previously shown to have a significant association with HNC: five autosomal dominant (*APC*, *BAP1*, *BRCA1*, *BRCA2*, and *RAD51B*), and one autosomal recessive (*MUTYH*).^23^ In our current study, the identified variants comprise 43 missense, 7-stop-gain, 3 frameshift, 2 in-frame substitution, and 2 intronic variants predicted to impact splicing (Figures 2 & 3). We compared the predicted damaging variants in these six genes between HNC cases and controls (Supplemental Table 1 and 2). We also compared the frequencies of in-frame indels, missense, and truncating variants in these six genes from our previous study, stratified by race and ethnicity.^23^ No significant associations between HNC and these genes were observed in the Nepalese population.

**Figure 2:**
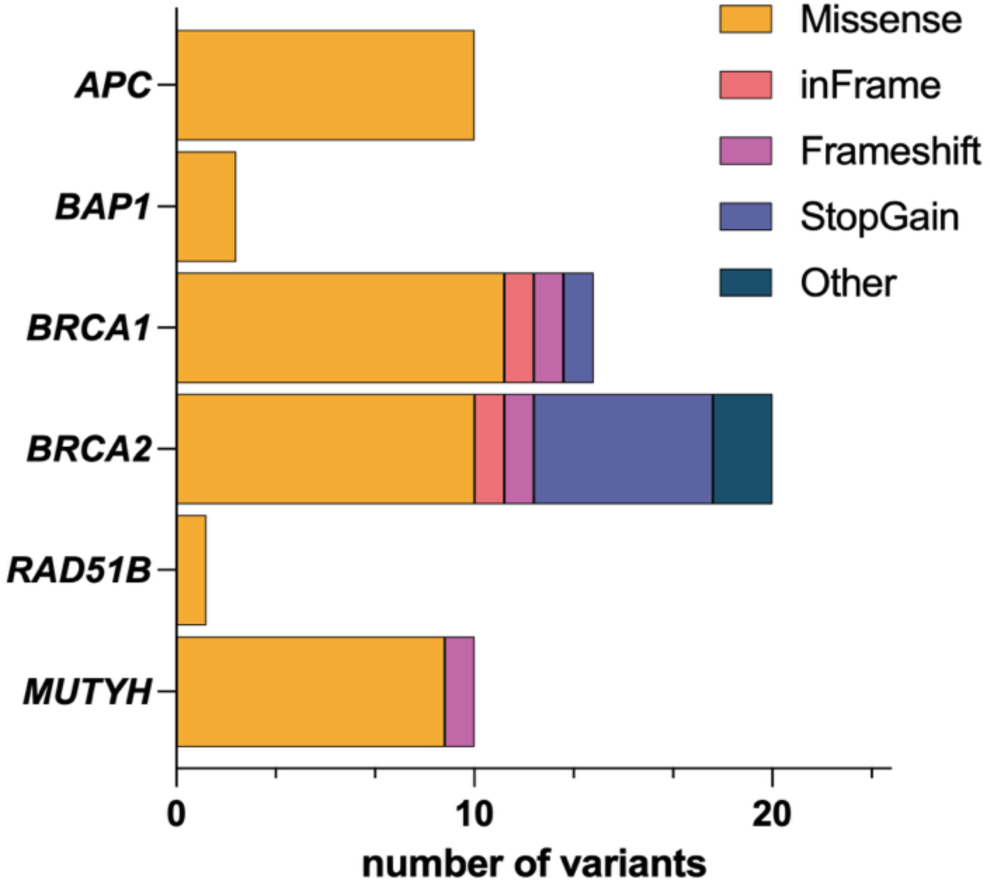
Distribution of variant types across cancer predisposition genes in the Nepal HNC cohort. Horizontal stacked bar chart showing the number and type of variants identified in *APC*, *BAP1*, *BRCA1*, *BRCA2*, *RAD51B*, and *MUTYH* from both cases and control. Variant categories include missense, in-frame insertion/deletion, frameshift, stop-gain, and other variant types, and are included independent of their annotated pathogenicity.

**Figure 3:**
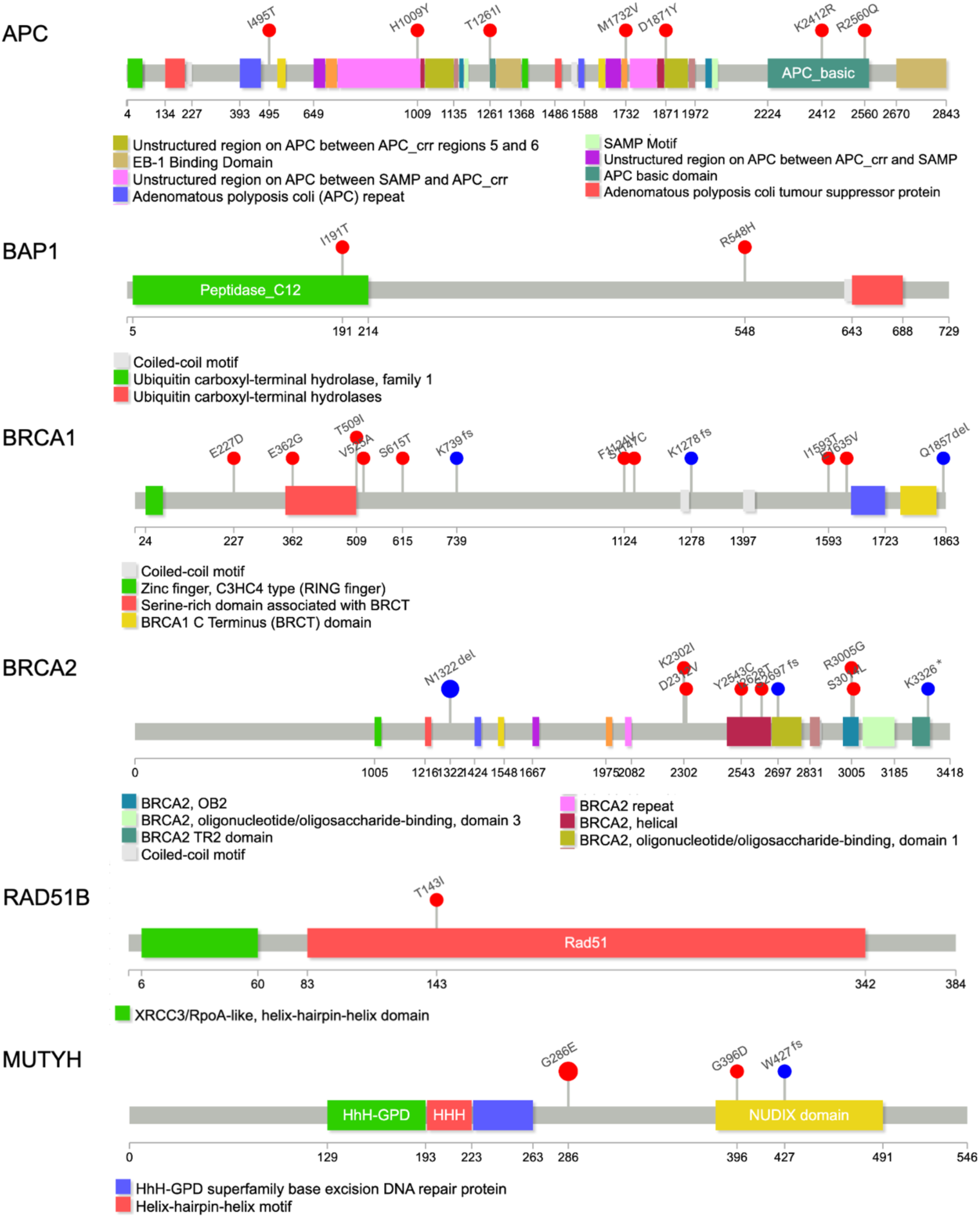
Lollipop plot showing the distribution of variants across the protein sequences of six cancer predisposition genes in the Nepal HNC cohort study. Red dots indicate missense variants. Blue dots indicate variants resulting in either frameshift (fs) or deletion (del). Numbers below each protein indicate amino acid position. Protein domains are highlighted. UniProt IDs used for protein annotation are APC (P25054), BAP1 (Q92560), BRCA1 (P38398), BRCA2 (P51587), RAD51B (O15315), and MUTYH (Q9UIF7).

Among APC carriers, all seven unique missense variants are classified as likely benign or benign. Two missense substitutions of unknown significance were observed in *BAP1*. In *BRCA1*, three truncating LoF variants were annotated as pathogenic, whereas the observed missense variants reside outside of the RING and BRCT domains and were annotated as likely benign. In *BRCA2*, a known benign stop-gain variant (p.K3326*) was detected in both cases and controls.

Other *BRCA2* variants included a pathogenic splice-site alteration and a frame-shift insertion. In *RAD51B*, a missense variant (p.T143I) with a high AlphaMissense score (+2) and predicted deleterious impact was found in a single case. In *MUTYH*, a monoallelic pathogenic frameshift variant p.W427fs was identified in one control, along with a recurrent monoallelic missense substitution p.G286E observed in 3 cases and 5 controls. In addition to a computational prediction of Pathogenic Supporting in this study, homozygous *MUTYH* p.G286E variants have been detected in patients with adenomatous polyposis, supporting its classification as pathogenic/likely pathogenic.^34,35^ Details of the identified variants, including AlphaMissense scores, BayesDel scores, and predicted functional impact, are provided in Supplemental Table 2.

## Discussion

Our study investigates HNC risk in Nepal, incorporating demographic, lifestyle, phenotypic, and genotypic data from a population with distinct exposure, habits, and socioeconomic conditions. There were significant differences between HNC cases and controls with cigarette smoking and alcohol consumption, which are established high-risk factors of HNC.^3–5,16,17^ Furthermore, this study reveals a significantly increased HNC burden with lower education level and income, underscoring the role of social determinants of cancer risk in this population. Importantly, our study replicates the association between chewing habits and increased HNC incidence.^17,22^ This reinforces the importance of using geographically and culturally diverse populations when evaluating HNC risk factors, as some chewing habits, such as betel quid, are particularly prevalent in Nepal.^36^

In the gene-based analysis, several genes, including *HSPG2*, *KRT6B*, *SLC4A2*, and *FREM1*, showed suggestive associations; however, none achieved statistical significance. These genes play roles in cytoskeletal structure, extracellular matrix organization, epithelial differentiation, and basement membrane formation, which are biological processes important for tumor invasion and metastasis.^37–41^ The candidate HNC predisposition genes identified in this study should be validated in a larger cohort. Similarly, although pathway-based analysis did not yield statistically significant results after multiple-testing correction, it identified several gene sets linked to extracellular structure organization, extracellular matrix remodeling, mammary gland development, transcription regulation, lipid metabolism, epigenetic regulation, and Cyclin D1 expression (Table 3). Some of these pathways have plausible roles in the pathogenesis and progression of HNC, thus providing a promising hypothesis for further study. In addition, it could be relevant in the context of environmental co-exposures common in Nepal, such as tobacco, alcohol, and betel quid.

The Phase II study identified nominally significant enrichment of pathogenic variants in six known cancer predisposition genes: *APC*, *BAP1*, *BRCA1*, *BRCA2*, *MUTYH*, and *RAD51B*. We further analyzed variants in these genes across both our Nepal cohort and the larger three-center case-control study, including Non-Hispanic White and East Asian cohorts, and did not observe statistically significant enrichment.^23^ Nonetheless, this comparison highlights the possibility of population-specific genetic contributions to HNC, emphasizing the importance of expanding genomic studies to underrepresented groups.

Clinical annotation of variants observed in our study was also performed following the ACMG protocol based on population, computational, and functional assays data for *APC*, *BAP1*, *BRCA1*, *BRCA2*, *MUTYH*, and *RAD51B*. The detection of clinically relevant variants collectively suggests that genomic heterogeneity exists in HNC. Among cancer susceptibility genes, most variants were missense substitutions. All *APC* missense variants identified were benign/likely benign, as they were located outside the mutation cluster region linked to high cancer risk. In contrast, several loss-of-function variants were detected in *BRCA1* and *BRCA2*, including in-frame deletions, frameshifts, stop-gain mutations, and splice-disrupting substitutions, most of which are annotated as pathogenic. Variants in *RAD51B* and *MUTYH* were mostly missense substitutions annotated with strong pathogenic or pathogenic-supporting evidence.

The most frequently observed pathogenic variant was MUTYH p.G286E, which has an overall MAF of 0.007053 in the study cohort. Notably, this pathogenic variant has been reported in East and South Asian populations at MAF of 0.0002450 and 0.0001757 respectively.^42^ Given that MUTYH-associated polyposis is a recessive condition, it is unsurprising to detect a recurrent pathogenic variant in controls. Indeed, 1 – 2% of non-Finnish Europeans are carriers of one of the well-studied pathogenic MUTYH founder mutations p.G396D or p.Y179C.^43^ Still, the number of p.G286E carriers in this Nepalese study cohort is significantly higher than reported for a general South Asian cohort (p-value 0.0033).^42^ This highlights the importance of expanding genetic studies to understudied populations, as there can be enrichment of specific variants, including pathogenic variants, in distinct subpopulations.

A key limitation of this study is the relatively small sample size for detecting pathogenic variants, which are inherently rare in the general population. The limited number of variant carriers reduced statistical power to detect effect sizes after correction for multiple testing in gene-based association tests. Another limitation is the potential for recall bias in the epidemiological data collected by retrospective questionnaires.

Importantly, this study provides a foundational genomic resource for HNC research in Nepal. It underscores two ideas: first, the importance expanding epidemiology studies to culturally-relevant risk factors and second, the need for ancestry-based cancer genomics. Ideally, this will entail combining epidemiological and genomic data in high-burden, underrepresented populations.

## Conclusion

This case-control study provides an integrated epidemiological and genomic analysis of HNC in Nepal and establishes behavioral risks showing a strong association of tobacco and alcohol in HNC. While no enrichment of genes and pathways was detected after correction for multiple testing, several clinically relevant variants were observed. Future research on HNC risk with a larger cohort in South Asians, especially in understudied and underrepresented groups, is warranted.

## Acknowledgements

The authors are greatly thankful to all the participants and study coordination team at BPKMCH. This work was partially funded by NIH 3P30 CA042014-26S6.

## Ethics Statement

The study was approved by the Institutional Review Boards (IRBs) at the University of Utah and the Nepal Health Research Council. All participants provided written informed consent.

## Data availability

The data of the current study are available upon request with appropriate institutional approvals, including IRB approval.

